# Diagnostic accuracy and acceptability of self- and health worker-collected tongue swabs for *Mycobacterium tuberculosis* complex detection in adults in South Africa

**DOI:** 10.64898/2026.07.04.26357275

**Authors:** Anura David, Yeonsoo Baik, Lesley Scott, Griffiths Kubeka, Adelaide Benoit, Lyndel Singh, Pedro da Silva, Wendy Stevens, Gregory P. Bisson, Salome Charalambous

## Abstract

Tongue swabs (TSs) are a non-invasive specimen type for the detection of *Mycobacterium tuberculosis* complex (MTBC) and can expand access to testing for individuals unable to produce sputum. This study evaluated the diagnostic performance and user acceptability of self-collected and health worker (HW)-collected tongue swabs using the Xpert MTB/RIF Ultra (Ultra) assay and assessed participant perspectives on self-collection.

In this prospective, cross-sectional study, symptomatic and asymptomatic adults under investigation for TB were enrolled from a high HIV prevalence setting. Each participant provided both a self-collected and a HW-collected TS, which were tested using Ultra. Ultra TS results were compared to liquid culture as the reference standard and sputum Ultra as a comparator. Participant perspectives on self-collection were captured via questionnaires.

Sensitivity on Ultra for both self- and HW-collected TSs was 68% (95% CI:51.9-81.9), compared to liquid culture. This sensitivity was significantly higher than that of sputum smear microscopy (46%, 95% CI: 30.7–62.6; McNemar’s *p = 0.003*). Tongue swab sensitivity was lower than sputum Ultra (80.5%; p<0.001) and decreased with low bacillary loads. Importantly, TSs enabled MTBC detection in six participants unable to produce sputum. Most participants (>90%) found self-collection instructions easy to follow, reporting high confidence and comfort, and trust in results from self-collected TSs.

This study demonstrates that self-collected TSs perform comparably to those collected by health workers for TB detection using Ultra and are both feasible and acceptable in a high TB/HIV burden setting. To maximize impact, clear training instructions and robust linkage to care remain critical priorities.

**Importance:** This study supports the use of tongue swabs (TSs) as a non-invasive alternative for tuberculosis diagnosis, particularly for individuals unable to produce sputum. When tested on the Xpert MTB/RIF Ultra assay, self-collected TSs performed comparably to health worker-collected swabs, yielding a 68% sensitivity (95% CI: 51.9–81.9) relative to liquid culture. This sensitivity was significantly higher than that of sputum smear microscopy (46%, 95% CI: 30.7–62.6; McNemar’s *p = 0.003*). Importantly, TSs successfully detected *Mycobacterium tuberculosis* complex in six patients who could not provide sputum, underscoring their clinical utility in expanding diagnostic access. Furthermore, high user acceptability (>90%) confirms that self-collection is both feasible and trusted by patients in high TB/HIV burden settings. To maximize real-world impact, implementing clear training instructions and establishing robust linkage to care, especially following negative results, remain critical programmatic priorities.

## Introduction

Tuberculosis (TB) remains a leading cause of morbidity and mortality globally (1). The burden of TB is disproportionately high in resource-limited settings, where access to timely and accurate diagnostic services is often constrained by infrastructure, human resources, and patient-related barriers (2). Conventional diagnostic approaches such as nucleic acid amplification tests (NAATs) and culture rely heavily on sputum-based testing, which poses challenges for individuals who are unable to expectorate (3). As a result, there is growing interest in additional non-sputum-based sampling methods that are less invasive, easier to collect, and more broadly accessible (4).

Tongue swab (TS) sampling has emerged as a viable approach for *Mycobacterium tuberculosis* complex (MTBC) detection using molecular assays such as Xpert® MTB/RIF Ultra (Ultra) (Cepheid, Sunnyvale, CA, USA) (5). Diagnosis of TB using TSs has also been recommended by the WHO, in individuals who cannot produce a sputum (6). Studies have demonstrated the feasibility of this method (7), but additional questions remain. Alongside optimizing specimen types, there is increasing interest in evaluating specimen collection methods that minimize reliance on health workers (HWs), thereby enhancing scalability and access, particularly in resource-limited or high-burden settings (8). Self-collection of specimens has been explored in various disease areas, including COVID-19 and other respiratory infections (9), as a strategy to decentralize care, reduce stigma, and enhance testing uptake. During the COVID-19 pandemic, Codsi et al. (10) found that supervised self-swabbing for TB was preferred over provider swabbing by many HWs due to perceived safety and reduced exposure risk. However, concerns remained about patients’ ability to perform self-collection accurately, highlighting the importance of patient education and simple, standardized collection instructions.

While tongue swabbing is gaining traction, no studies to date, that we are aware of, have directly compared the diagnostic performance of self-collected versus HW-collected tongue swabs for MTBC detection. The comparative accuracy, acceptability, and operational implications of these collection methods are critical to assess for informing broader implementation of TSs as a diagnostic tool, particularly in community-based or resource-limited settings where HW availability may be constrained.

In this study, we compared the diagnostic performance of self-collected and HW-collected TSs for MTBC detection using the Ultra assay. In addition, the study assessed participants’ preferences, confidence, ease of use of self-collection as well as opinions on self-collection strategies through completion of a survey.

## Materials and Methods

### Study design and participants

Participant recruitment for this study was conducted in two phases. Phase one was conducted from 23 January to 06 April 2023 at five healthcare facilities (HCFs) in the Ekurhuleni District, Gauteng, South Africa: Germiston Clinic, Goba Clinic, Voslorus Poly Clinic, Phola Park Clinic, and Ramokonopi Clinic. To incorporate emerging evidence on optimized swab processing protocols, a second phase of the study was implemented. Phase two took place from 25 March to 08 May 2024 at these same five HCFs, with the addition of three more: J Dumane Clinic, Rondebult Clinic, and Wannenburg Clinic.

Clinic staff referred asymptomatic individuals undergoing TB investigation as well as symptomatic adults (aged ≥18 years) with presumed TB as determined by local HWs to the study team for potential enrolment. To be eligible, participants needed to provide informed consent, be willing to provide an additional sputum sample, agree to self-collect a TS, and not be on TB treatment. Following consent, data on symptoms, demographics, medical history, and current health status were collected using a standardized reporting tool.

Study specimens were collected during a single visit (Figure 1). For study purposes, a sputum and two TSs (one self-collected and one HW-collected) were collected. For phase one, a short-shaft (4 cm) spun polyester XpressCollect^TM^ swab (Steripack, USA) (n=314) was used. While this swab was initially selected to evaluate its viability for TB diagnosis, preliminary data analysis revealed poor participant acceptability, primarily due to the short shaft length complicating usability. Consequently, for phase two, the protocol was adapted to utilize a longer-shaft (15 cm) Copan FLOQSwab® (Copan Italia S.p.A.) (n=355) with a 30 mm breakpoint. Swab collection involved swabbing the back and top of the tongue in back-front and left-right motions for 10 seconds while rotating the swab and avoiding a gag reflex.

**Figure 1:**
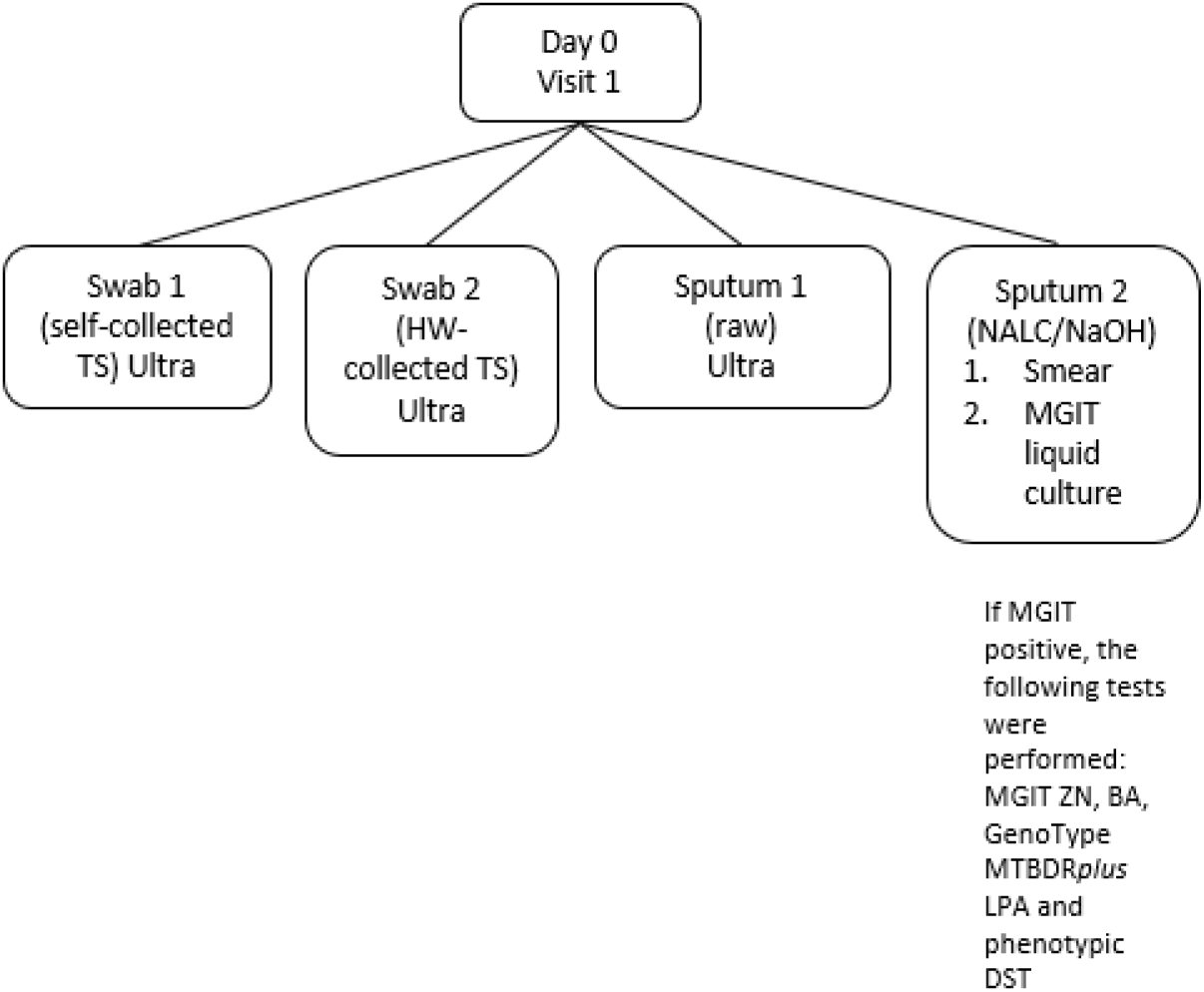
Specimen Collection and Testing Overview. The schematic outlines the collection and processing pathways for the two sputum specimens and two tongue swabs obtained from each participant. Ultra, Xpert MTB/RIF Ultra; TS, tongue swab; HW, health worker; NaOH/NALC, sodium hydroxide N-acetyl-L-cysteine, MGIT, mycobacterial growth indicator tube; ZN, Ziehl-Neelsen smear, BA, blood agar; LPA, line-probe assay

Participants first performed self-collection of TSs guided by written instructions (Supplementary figure 1) while being observed by a HW, who did not assist or intervene. This was followed by TS collection performed by the HW. Afterwards, participants were asked a series of questions about their self-collection experience, health-seeking behaviours, and their opinions on the acceptability and feasibility of self-collection.

Sputum 1 was sent for standard-of-care testing to the National Health Laboratory Services (NHLS). Sputum 2 and both TSs were transported at 2-8°C to the research laboratory in Braamfontein, Johannesburg for testing.

### Laboratory testing

Sputum were processed using N-acetyl-L-cysteine/Sodium hydroxide (NALC/NaOH) for smear and Mycobacterial Growth Indicator Tube (MGIT) (Becton Dickinson, Sparks, MD, USA) liquid culture using adapted procedures from the MGIT procedure manual (11) and the BACTEC MGIT 960 System User’s Manual. Result return was performed by laboratory staff as per the current National Tuberculosis TB Management Guidelines (12). Tongue swabs in phase one were pre-processed using phosphate buffer (PB; Media Mage, Johannesburg, SA) before testing on the Ultra assay. Briefly, 2.3 mL of PB was added to the TS, vortexed for 20 seconds and incubated for 5 minutes. A total volume of 1.5 mL of the resulting liquid was then tested on Ultra. For phase 2, a diluted sample reagent (SR) buffer protocol for TS testing on Ultra was followed (13).

### Outcomes and statistical analysis

The study evaluated the diagnostic accuracy of self-collected and HW-collected TSs tested with Ultra for detecting MTBC. Liquid culture and phenotypic drug-susceptibility testing (pDST) (performed using the MGIT960 SIRE kit [Becton Dickinson, Sparks]) were used as reference standards for MTBC detection and for rifampicin (RIF) resistance detection. Data analysis included calculation of sensitivity, specificity, positive predictive value, and negative predictive value for MTBC detection and concordance for RIF resistance detection, with 95% CIs calculated using the Wilson score method. Ultra sputum was used as a comparator assay. McNemar’s test was used to compare the sensitivity of TS Ultra against both sputum Ultra and sputum smear microscopy. Two-sided *p*-values less than 0.05 were considered statistically significant.

For sample size calculation, a 10% prevalence of TB among tested individuals was assumed in outpatient clinic settings in South Africa, and a minimum of 660 samples was required to achieve at least 80% power, 10% effect size, and 95% accuracy. In determining the sample size, allowance was made for patients who do not meet the inclusion criteria. To determine the performance of the Ultra TS assay, only specimens that generated valid results across all assays (Ultra and MGIT) were included in the statistical analysis.

### Ethics statement

The University of the Witwatersrand Human Research Ethics Committee (#220411) and the University of Pennsylvania School of Medicine (#851244) gave ethical approval for this work.

## Results

### Participant characteristics

In this prospective, cross-sectional study, 669 participants provided informed consent and were enrolled (Figure 2). Among these, sputum for smear and culture was unavailable for 47 participants; TSs successfully detected MTBC in 6/47 (13%) individuals (two on both swab types, and four on either the HW- or self-collected swab). Where valid results were available for each test, TB was bacteriologically confirmed in 8% using culture only, 6% using Ultra sputum only and 5% using Ultra TS only. A total of 149 participants (inclusive of the 47 without sputum) were excluded due to unsuccessful laboratory results (n=93) from Ultra sputum and/or TS or liquid culture, or due to missing or rejected specimens (n=56). Thus, 520 participants were included in the statistical analysis.

**Figure 2:**
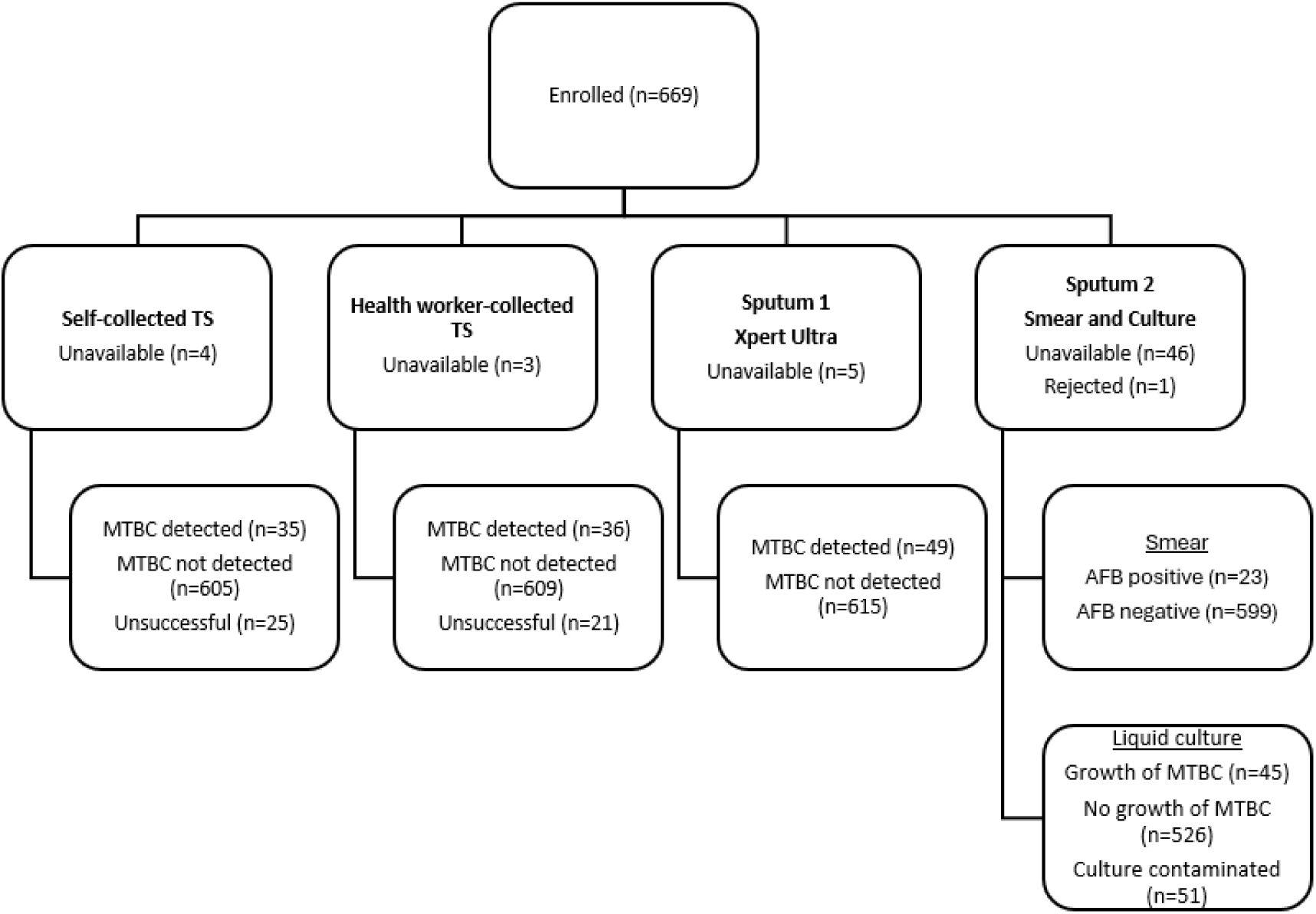
Flow diagram of participant enrolment, specimen distribution, and Xpert Ultra and liquid culture results. Enrolled individuals (N = 669) provided two sputum specimens and two tongue swabs. Final diagnostic classifications (MTBC detected, not detected, missing, or unsuccessful) are provided for each individual test arm. TS, tongue swab; MTBC, Mycobacterium tuberculosis complex

Among the 520 participants, 54% were male with an average age of 42 years (Table 1). A total of 44% were people with HIV (PHIV) with 6% of unknown status. Previous TB was reported by 4% of participants. Cough was the most frequently reported symptom among participants while 12/514 (2.3%) were asymptomatic. Forty-eight participants reported underlying health conditions and up to 40% of participants reported additional risk factors for TB. Bacteriological confirmation of TB using liquid culture was seen in 41/520 (8%) of participants.

**Table 1:** Characteristics of study participants included in the statistical analysis.

| **Characteristic** |  |
| --- | --- |
| *Demographics (n=520)* |  |
| Age, mean (range), years | 42 (18-84) |
| Male sex, n (%) | 281 (54) |
| *HIV-related information* |  |
| PHIV, n/N (%) | 229/520 (44) |
| HIV-negative, n/N (%) | 262/520 (50) |
| Status unknown, n/N (%) | 29/520 (6) |
| On ARVs, n/N (%) | 200/229 (87) |
| *TB History* |  |
| Previously diagnosed with TB, n/N (%) | 21/513 (4) |
| *Clinical signs and symptoms of TB at presentation* |  |
| Unexplained weight loss, n/N (%) | 177/493 (36) |
| Nights sweats, n/N (%) | 251/512 (49) |
| Fever, n/N (%) | 243/511 (48) |
| Cough (any duration), n/N (%) | 479/514 (93) |
| Haemoptysis, n/N (%) | 38/520 (7) |
| *Additional TB risk factors* |  |
| *Underlying health conditions, n/N (%) | 48/520 (9) |
| Known TB contact, n/N (%) | 98/483 (20) |
| Alcohol consumption (more than once a week), n/N (%) | 111/227 (40) |
| Smoker, n/N (%) | 163/509 (31) |
| *Bacteriological confirmation (n=520)* |  |
| Smear and culture positive, n (%) | 19 (4) |
| Smear negative and culture positive, n (%) | 22 (4) |
| Smear and culture negative, n (%) | 478 (92) |
| Smear positive and culture negative, n (%) | 1 (<1) |

ARV, antiretroviral; PHIV, people with HIV; \*Underlying health conditions include diabetes, asthma, bronchitis, high blood pressure and angina

### Diagnostic performance of Ultra tongue swabs compared to liquid culture

Among culture-confirmed TB cases, the sensitivity of both self-collected and HW-collected TS on Ultra was 68.3% (28/41; 95% CI: 51.9–81.9). Specificity was similarly high for both methods: 99.8% (95% CI: 98.8–100) for self-collected TSs and 99.4% (95% CI: 98.2–99.9) for HW-collected TSs (Table 2). Ultra TS sensitivity was significantly higher than that of sputum smear microscopy (46%, 95% CI: 30.7–62.6; McNemar’s *p = 0.003*). Tongue swab sensitivity was lower than that of sputum Ultra (80.5%; 33/41, McNemar’s *p<0.001*).

**Table 2:** Diagnostic performance of smear microscopy and Xpert Ultra assays (sputum and tongue swab) compared with liquid culture for MTBC detection.

| Variable | Smear microscopy | n/N | Xpert MTB/RIF Ultra (sputum) | n/N | Xpert MTB/RIF Ultra (self-collected TS) | n/N | Xpert MTB/RIF Ultra (HW-collected TS) | n/N |
| --- | --- | --- | --- | --- | --- | --- | --- | --- |
| *All (including trace) (n=520)* |  |  |  |  |  |  |  |  |
| Sensitivity, % (95% CI) | 46.3 (30.7-62.6) | 19/41 | 80.5 (65.1–91.2) | 33/41 | 68.3 (51.9-81.9) | 28/41 | 68.3 (51.9-81.9) | 28/41 |
| Specificity, % (95% CI) | 99.8 (98.8-100) | 478/479 | 98.1 (96.5–99.1) | 470/479 | 99.8 (98.8-100) | 478/479 | 99.4 (98.2-99.9) | 476/479 |
| PPV, % (95% CI) | 95.0 (75.1-99.9) | 19/20 | 78.6 (63.2–89.7) | 33/42 | 96.6 (82.2-99.9) | 28/29 | 90.3 (74.2-98.0) | 28/31 |
| NPV, % (95% CI) | 95.6 (93.4–97.2) | 478/500 | 98.3 (96.7–99.3) | 470/478 | 97.4 (95.5-98.6) | 478/491 | 97.3 (95.5-98.6) | 476/489 |
| *Specimens from HIV-positive individuals (n=229)* |  |  |  |  |  |  |  |  |
| Sensitivity, % (95% CI) | 55.0 (31.5-76.9) | 11/20 | 95.0 (75.1–99.9) | 19/20 | 80.0 (56.3-94.3) | 16/20 | 85.0 (62.1-96.8) | 17/20 |
| Specificity, % (95% CI) | 100 (98.3-100) | 209/209 | 95.7 (92.0–98.0) | 200/209 | 99.5 (97.4-100) | 208/209 | 99.5 (97.4-100) | 208/209 |
| PPV, % (95% CI) | 100 (71.5–100) | 11/11 | 67.9 (47.6–84.1) | 19/28 | 94.1 (71.3-99.9) | 16/17 | 94.4 (72.7-99.9) | 17/18 |
| NPV, % (95% CI) | 95.9 (92.3–98.1) | 209/218 | 99.5 (97.3–100) | 200/201 | 98.1 (95.2-99.5) | 208/212 | 98.6 (95.9-99.7) | 208/211 |
| *Specimens from HIV-negative individuals (n=262)* |  |  |  |  |  |  |  |  |
| Sensitivity, % (95% CI) | 36.8 (16.3-61.6) | 7/19 | 63.2 (38.4–83.7) | 12/19 | 57.9 (33.5-79.7) | 11/19 | 52.6 (28.92-75.6) | 10/19 |
| Specificity, % (95% CI) | 99.6 (97.7-100) | 242/243 | 100 (98.5–100) | 243/243 | 100 (98.5-100) | 243/243 | 99.2 (97.1-99.9) | 241/243 |
| PPV, % (95% CI) | 87.5 (47.3-99.7) | 7/8 | 100 (73.5–100) | 12/12 | 100 (71.5-100) | 11/11 | 83.3 (51.6-97.9) | 10/12 |
| NPV, % (95% CI) | 95.3 (91.9-97.5) | 242/254 | 97.2 (94.3–98.9) | 243/250 | 96.8 (93.8–98.6) | 243/251 | 96.4 (93.3–98.3) | 241/250 |
| *Smear microscopy negative specimens (n=500)* |  |  |  |  |  |  |  |  |
| Sensitivity, % (95% CI) | n/a |  | 68.2 (45.1-86.1) | 15/22 | 45.5 (24.4-67.8) | 10/12 | 40.9 (20.7-63.6) | 9/22 |
| Specificity, % (95% CI) |  |  | 98.1 (96.5–99.1) | 469/478 | 99.8 (98.8-100) | 477/478 | 99.4 (98.2-99.9) | 475/478 |
| PPV, % (95% CI) |  |  | 62.5 (40.6–81.2) | 15/24 | 90.9 (58.7-99.8) | 10/11 | 75.0 (42.8-94.5) | 9/12 |
| NPV, % (95% CI) |  |  | 98.5 (97.0–99.4) | 469/476 | 97.5 (95.8-98.7) | 477/489 | 97.3 (95.5-98.6) | 475/488 |

95% CI, 95% confidence intervals; TS, tongue swab; PPV, positive predictive value; NPV, negative predictive value

### Diagnostic performance of Ultra tongue swabs compared to Ultra sputum

Both self-collected and HW-collected TSs showed slightly lower sensitivity (66.7%; 95% CI: 50.5–80.4) and comparable specificity (99.4%; 95% CI: 98.2–99.9) when compared to Ultra sputum, rather than to liquid culture. TS was able to detect MTBC across the semi-quantitative range, with some variability (Table 3).

**Table 3:** Comparison of Ultra sputum semi-quantitative results with Ultra TS.

| Ultra sputum semi-quantitative result | Ultra TS result (self-collected) |  | Ultra TS result (HW-collected) |  |
| --- | --- | --- | --- | --- |
|  | n/N | Percent detection | n/N | Percent detection |
| High | 16/21 | 76.2 | 17/21 | 80.9 |
| Medium | 1/1 | 100 | 1/1 | 100 |
| Low | 3/3 | 100 | 3/3 | 100 |
| Very low | 1/4 | 25 | 0/4 | 0 |
| Trace | 3/6 | 50 | 3/6 | 50 |

Ultra refers to the Xpert MTB/RIF Ultra assay, HW, health worker; TS, tongue swab;

### RIF resistance detection using the tongue swab

No RIF resistance was detected on participants enrolled on this study. Where valid Ultra TS RIF results were available, there was 100% concordance with pDST.

### Errors on Ultra tongue swabs

Of all TSs tested on the Ultra assay, 46/1331 (4%) demonstrated errors (Figure 3).

**Figure 3:**
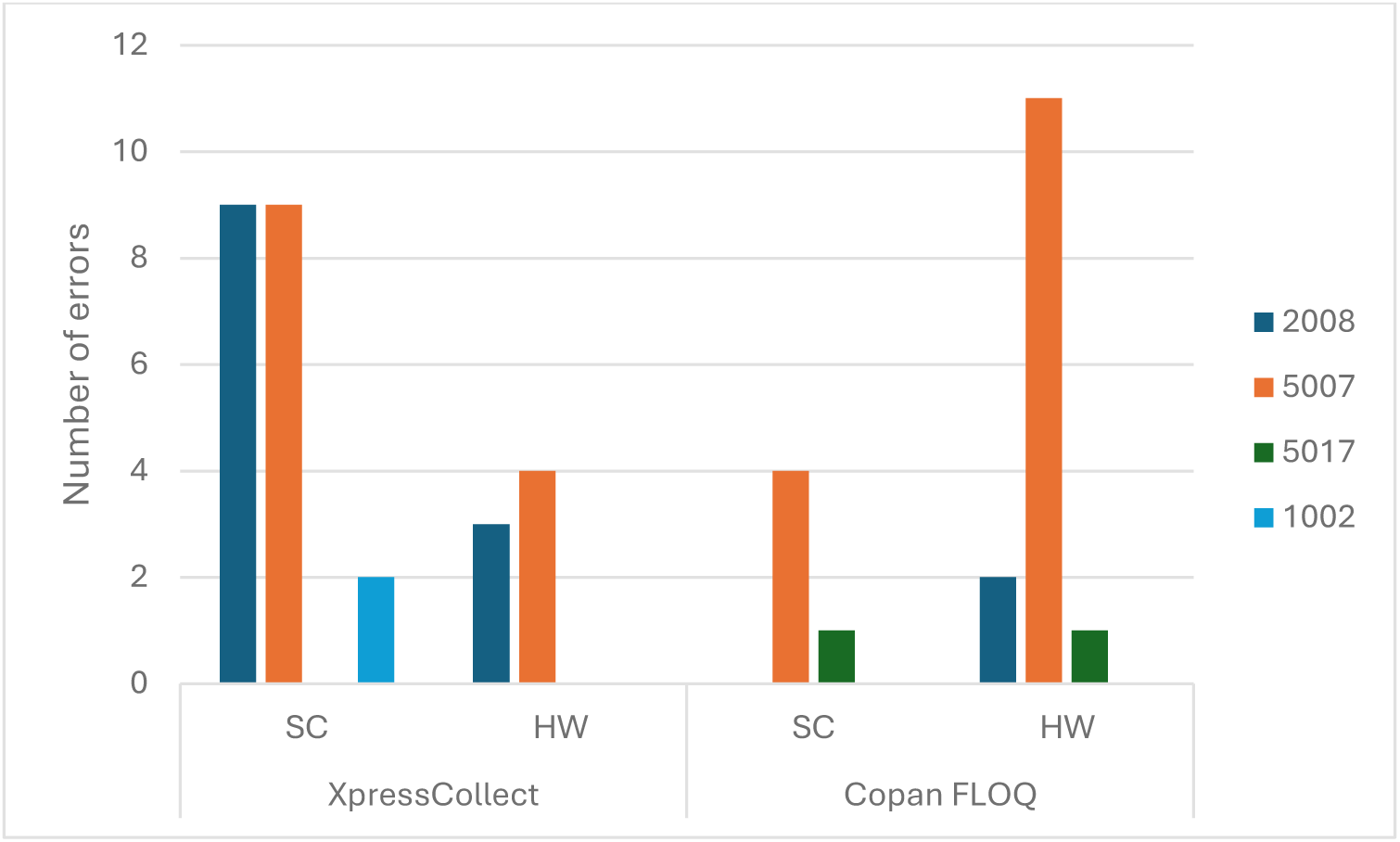
Frequency and error codes observed on the Ultra using tongue swabs. The figure displays the total number and distribution of error codes during testing of both tongue swab types using the Ultra assay. Error categories are based on assay-defined codes and include error 1002: temperature or heater failure; error 2008: abnormal pressure detected; error 5007 and 5017: probe check failed; SC, self-collection; HW, health worker

### Observation of self-collection by the HW

Among participants observed during self-collection, hand hygiene was performed by 514/519 (99%), and 517/518 (99%) avoided touching the swab head. A correct swabbing duration of 10 seconds was performed by 381/519 (73%). After specimen collection, 513/517 (99%) participants inserted the swab and closed the tube correctly.

### Health seeking behavior of participants

Among survey respondents, 432/509 (85%) indicated they would be willing to perform self-collection when feeling ill. Currently, 324/518 (63%) reported that they typically seek medical care when unwell. When asked about preferences if self-collection were routinely available, 266/513 (52%) stated they would prefer to visit a clinic, whereas 240/513 (47%) would opt for the self-collection option. If a self-collected TS tested positive, 406/519 (78%) indicated they would seek care. In contrast, only 119/516 (23%) would seek care following a negative result, while 299/516 (58%) reported they would rarely or never pursue care. Additionally, 424/518 (82%) of participants believed that results from a self-collected TS would be as accurate as those collected by a HW.

### Participants responses to tongue swab collection

Most participants (>90%) found TS self-collection instructions easy to follow, reported that self-collection was easy and were comfortable and confident with self-collection. Of the 518 respondents, 223 (43%) didn’t have a preference between HW or self-collection, 183 (35%) preferred HW collection and 112 (21%) preferred self-collection. Participants using XpressCollect swabs reported difficulty with collection due to the short swab shaft.

### Participant preferences on self-collection

Participants preferences for receipt of self-collection kits, location of self-sampling and return of self-collected swabs are indicated in Figure 4.

**Figure 4:**
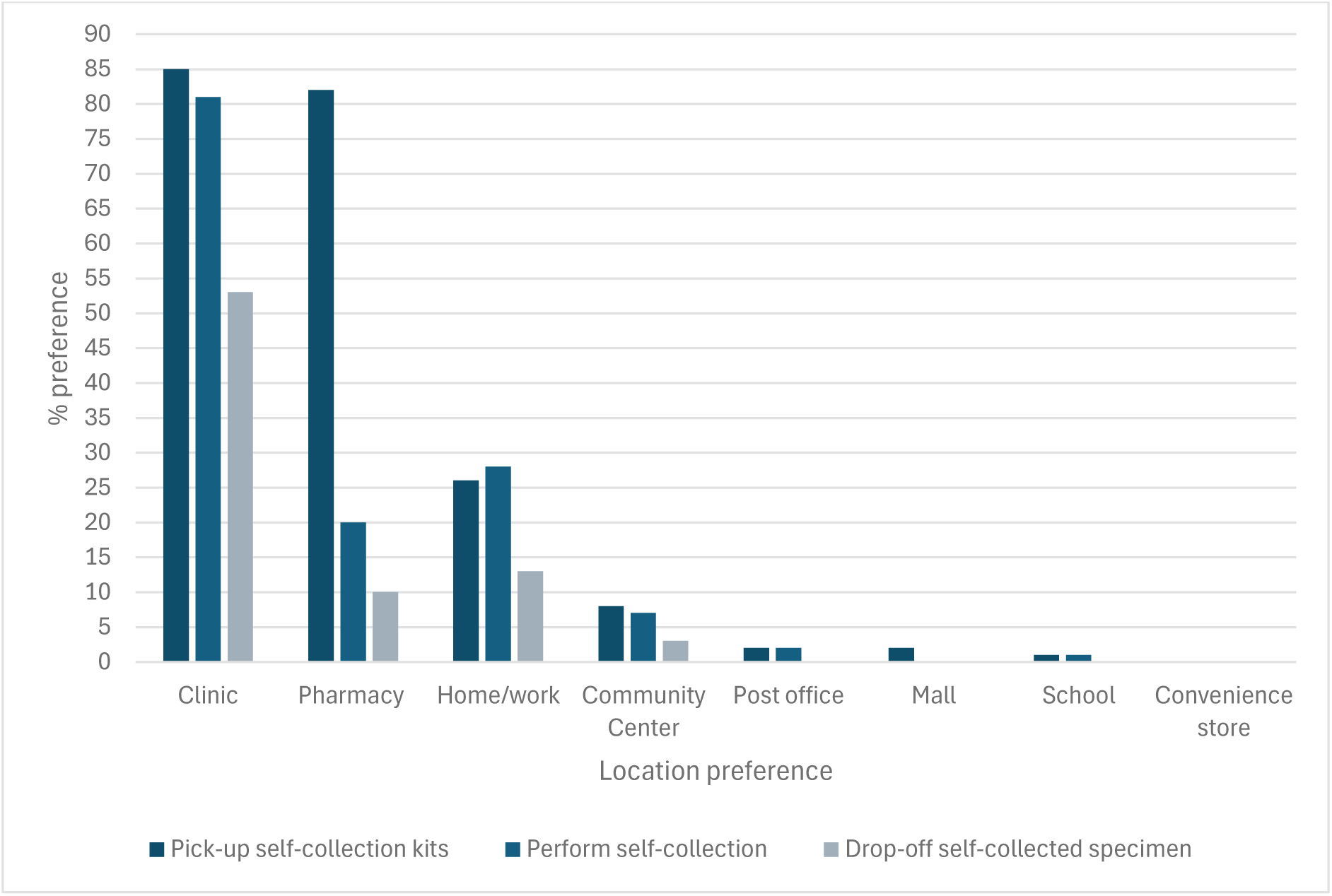
Participant preferences for self-sampling kit collection, self-sampling location and self-collection specimen return. Participants selected from predefined options including clinics, pharmacies, home/work, community centres, schools, convenience stores, malls, and post offices. The majority preferred clinics and pharmacies across all three categories, while home/work and community centres were moderately preferred and post offices, malls, schools and convenience stores were least favoured.

## Discussion

In this study which compared the diagnostic performance of self-collected and HW-collected TSs for TB, self-collected swabs demonstrated comparable performance to HW-collected TSs using the Ultra assay. Both methods yielded moderate sensitivity and high specificity relative to liquid culture. Participants generally reported self-collection was easy, acceptable, and feasible.

The diagnostic performance observed aligns with previous evaluations of oral and TS testing for TB diagnosis (5, 7). While TS sensitivity was lower than sputum Ultra, it surpassed sputum smear microscopy sensitivity in this population. Notably, TSs enabled MTBC detection in participants unable to provide sputum, highlighting the value of this specimen type in this population. These findings support the use of TSs as a complementary diagnostic specimen rather than a replacement for sputum-based testing.

Several methodological factors may have influenced the diagnostic performance observed. Two different swab types and two pre-processing protocols were used across study phases, possibly contributing to variability in assay results. Previous research shows swab design affects the amount of oral biomass collected (14). Participants were instructed to swab for ten seconds, while current consensus recommends 30 seconds (15). About one-quarter of participants did not complete the recommended duration during observed self-collection, which may have reduced organism recovery and sensitivity. Future studies should evaluate standardized swab materials, optimized processing methods, and adherence to updated protocols to better assess the diagnostic potential of TSs testing.

Participants unable to provide sputum, who may benefit most from non-sputum-based testing, could not be fully included in the primary diagnostic accuracy analysis, due to missing reference standard results. Consequently, the evaluated population may not fully represent the intended target group for TS testing. Notably, MTBC was detected by TS testing in six participants without sputum, highlighting the clinical value of TS sampling in sputum-scarce individuals. Use of a liquid culture reference standard illustrates a broader challenge in evaluating non-sputum TB diagnostics, as conventional microbiological reference standards depend on sputum availability.

The overall unsuccessful test rate for TSs on Ultra was low; and assay errors were more frequent with the XpressCollect swab than the Copan FLOQ swab. The most common error, code 5007, is typically linked to viscous specimens, insufficient sample volume, or processing issues. These findings highlight the importance of using optimized specimen processing protocols and selecting swab types compatible with molecular testing workflows.

The small number of culture-confirmed TB cases among PHIV warrants cautious interpretation of these subgroup findings. Improved sensitivity on PHIV was reported in a previous study evaluating oral swabs for TB diagnosis in a high HIV burden setting (16), but larger studies are needed to clarify the relationship between HIV status, bacillary burden, and TS performance. Additionally, the moderate sensitivity observed among smear-negative participants highlights the persistent challenges of detecting paucibacillary disease using TSs.

Participant responses indicated high acceptability of self-collection. Most reported the instructions were easy to follow and felt comfortable and confident performing the procedure. Many believed self-collected TSs would yield results comparable to HW-collected specimens. Preferences varied, with some favoring HW collection, others preferring self-collection, and some expressing no preference. Clinics and pharmacies were the most preferred locations for obtaining and returning self-collection kits, suggesting participants value convenience while still wanting some engagement and support from the healthcare system during testing.

The survey findings highlighted important considerations for implementation. While most participants said they would seek care after a positive self-collected test result, substantially fewer, including those with symptoms, said they would seek medical care after a negative result. This raises concerns about potential false reassurance and delayed healthcare seeking if TS self-collection is used in TB screening. Clear patient education, counseling, and robust linkage-to-care systems will be essential for future self-collection programs. Lessons from HIV self-testing programs in South Africa suggest digital health interventions, such as mobile applications, SMS reminders, and electronic result delivery, may support correct specimen collection, facilitate follow-up, and improve linkage to care (17–19).

This study has limitations such as the use of different swab types and laboratory processing methods across study phases. Observation during self-collection may have influenced acceptability responses due to social desirability bias. Finally, the number of rifampicin-resistant cases was too small to robustly evaluate resistance detection using TSs.

In conclusion, self-collected TSs demonstrated comparable diagnostic performance to HW–collected TSs for MTBC detection using the Ultra assay and were generally well accepted by participants. Although TS sensitivity was lower than that of sputum tested by Ultra, this specimen type provides an important alternative for individuals unable to produce sputum and could facilitate decentralised or community-based TB case-finding strategies. Self-collection also represents an attractive approach for emerging swab-based near point-of-care technologies recommended by the WHO (6). Further research is needed to evaluate implementation in unsupervised settings and to assess strategies for ensuring effective linkage to care following self-collection.

## Acknowledgments

We thank the study participants; the Johannesburg Health District and the and the Gauteng Department of Health for their support and collaboration on this study; clinical research staff, Wits DIH TB laboratory staff.

## Data availability statement

All data produced in the present study are available upon reasonable request to the authors.

## Author contributions

Conceptualization, Yeonsoo Baik; Data curation, Anura David, Griffiths Kubeka, Yeonsoo Baik; Formal analysis, Anura David; Funding acquisition, Gregory P Bisson, Yeonsoo Baik, Lesley Scott, Wendy Stevens; Investigation, Anura David, Griffiths Kubeka, Lyndel Singh; Methodology, Anura David, Yeonsoo Baik; Project administration, Griffiths Kubeka, Anura David; Supervision, Salome Charalambous, Gregory P Bisson, Lesley Scott; Validation, Anura David; Visualization, Yeonsoo Baik, Gregory P Bisson, Salome Charalambous; Writing—original draft, Anura David; Writing—review and editing, Anura David, Gregory P. Bisson, Salome Charalambous, Lesley Scott, Griffiths Kubeka, Lyndel Singh, Pedro da Silva, Wendy Stevens, Yeonsoo Baik

All authors have read and agreed to the published version of the manuscript.

## References

1. Global Tuberculosis Report 2025. World Health Organisation; 2025 https://www.who.int/teams/global-programme-on-tuberculosis-and-lung-health/tb-reports/global-tuberculosis-report-2025.

2. Cioboata R, Balteanu MA, Osman A, Vlasceanu SG, Zlatian OM, Mitroi DM, et al. Coinfections in Tuberculosis in Low- and Middle-Income Countries: Epidemiology, Clinical Implications, Diagnostic Challenges, and Management Strategies—A Narrative Review. J Clin Med. 2025;14(7):2154.

3. WHO. Target product profiles for tuberculosis diagnosis and detection of drug resistance; 2024 https://www.who.int/publications/i/item/9789240097698.

4. WHO. High-Priority Target Product Profiles for New Tuberculosis Diagnostics: Report of a Consensus Meeting Geneva, Switzerland 2014 https://www.who.int/tb/publications/tpp_report/en/.

5. Wood RC, Luabeya AK, Dragovich RB, Olson AM, Lochner KA, Weigel KM, et al. Tongue swab testing on two automated tuberculosis diagnostic platforms, Cepheid Xpert® MTB/RIF Ultra and Molbio Truenat® MTB Ultima. J Clin Microbiol. 2024;10(4).

6. World Health Organization. WHO issues new recommendations on near point-of-care tests for TB diagnosis, including tongue swabs. Geneva: WHO; 2026. https://www.who.int/teams/global-programme-on-tuberculosis-and-lung-health/diagnosis-treatment/npoc-tongue-swabs-and-sputum-pooling-for-tb

7. Church EC, Steingart KR, Cangelosi GA, Ruhwald M, Kohli M, Shapiro AE. Oral swabs with a rapid molecular diagnostic test for pulmonary tuberculosis in adults and children: a systematic review. Lancet Glob Health. 2024;12:e45–54.

8. Drain PK, Gardiner J, Hannah H, Broger T, Dheda K, Fielding K, et al. Guidance for studies evaluating the accuracy of biomarker-based nonsputum tests to diagnose tuberculosis. J Infect Dis. 2019;8(220):S108–S15.

9. Cockerill FR, Wohlgemuth JG, J. R, Sabol CE, Kapoor H, Dlott JS, et al. Evolution of Specimen Self-Collection in the COVID-19 Era: Implications for Population Health Management of Infectious Disease. Popul Health Manag. 2021;24(1):S26–S34.

10. Codsi R, Errett NA, Luabeya AK, Van As D, Hatherill M, Shapiro AE, et al. Preferences of healthcare workers using tongue swabs for tuberculosis diagnosis during COVID-19. PLOS Glob Public Health. 2023;3(9).

11. FIND. MGIT Manual: Manual for BACTEC MGIT 960 TB System. Geneva: Foundation for Innovative New Diagnostics; 2006. https://www.finddx.org/wp-content/uploads/2023/02/20061101_rep_mgit_manual_FV_EN.pdf

12. South African National Department of Health. National Tuberculosis Management Guidelines. https://knowledgehub.health.gov.za/elibrary/national-tuberculosis-management-guidelines.

13. Ahls C, David A, Chilambi GS, Cattamanchi A, de Vos M, Heard K, et al. Xpert MTB/RIF Ultra testing from tongue swabs - Diluted SR method. 2024 https://www.protocols.io/view/xpert-mtb-rif-ultra-testing-from-tongue-swabs-dilu-14egn69nyl5d/v1.

14. Wood RC, Andama A, Hermansky G, Burkot S, Asege L, Job M, et al. Characterization of oral swab samples for diagnosis of pulmonary tuberculosis. PLOS ONE. 2021;16(5):e0251422.

15. Andama A, Steadman AE, Ahls C, Cangelosi GA, David A, de Vos M, et al. Consensus standard operating procedure for collection of tongue swabs for TB diagnostics 2024 https://www.protocols.io/view/consensus-standard-operating-procedure-for-collect-kxygxyw54l8j/v1.

16. LaCourse SM, Seko E, Wood R, Bundi W, Ouma GS, Agaya J, et al. Diagnostic performance of oral swabs for non-sputum based TB diagnosis in a TB/HIV endemic setting. PLoS ONE. 2022;17(1):e0262123.

17. Mshweshwe-Pakela NT, Mabuto T, Shankland L, Fischer A, Tsukudu D, Hoffmann CJ. Digitally supported HIV self-testing increases facility-based HIV testing capacity in Ekurhuleni, South Africa. South Afr J HIV Med. 2022;23(1):1352.

18. Gous N, Fischer AE, Rhagnath N, Phatsoane M, Majam M, Lalla-Edward ST. Evaluation of a mobile application to support HIV self-testing in Johannesburg, South Africa. 2020. 2020;21(1).

19. World Health Organization. Digital health for the End TB Strategy: an agenda for action. Geneva: WHO; 2015. https://www.who.int/publications/i/item/WHO-HTM-TB-2015.21.

