## Supplementary Figure 1 for "Diagnostic accuracy and acceptability of self- and health worker-collected tongue swabs for *Mycobacterium tuberculosis* complex detection in adults in South Africa"

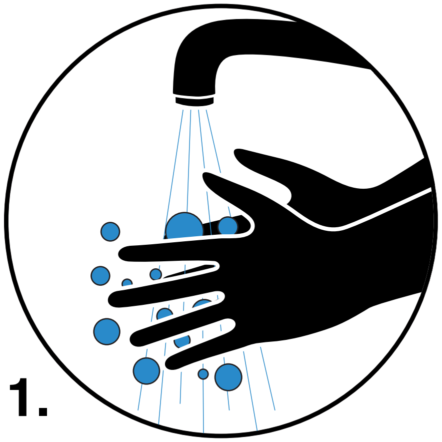

1\.

Wash hands

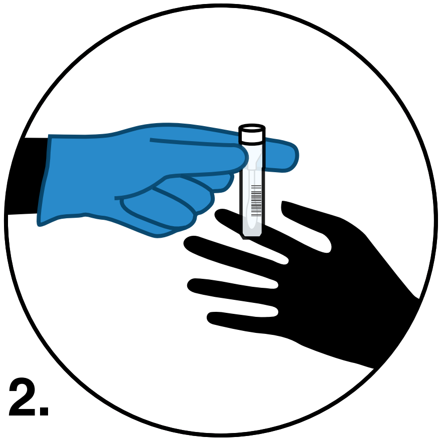

2\.

Check your information on the tube

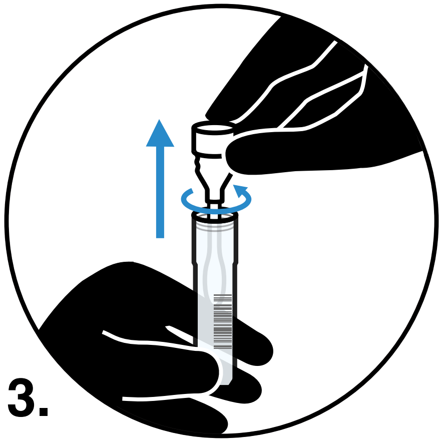

3\.

Open the tub cap

DO NOT TOUCH SWAB WITH HANDS

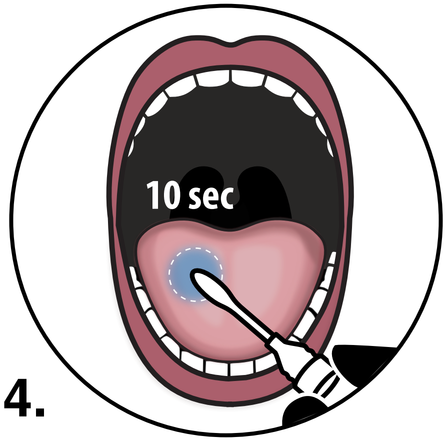


4\.

Swab the tongue for 10 seconds

DO NOT TOUCH SWAB WITH HANDS

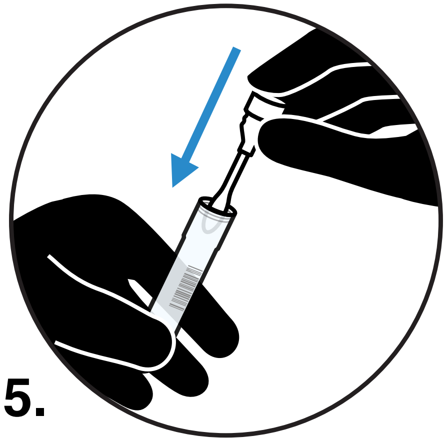

5\.

Place swab in tube and seal the tube cap

DO NOT TOUCH SWAB WITH HANDS

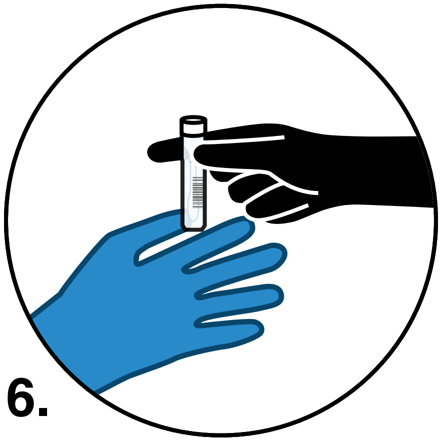

6\.

Submit the tube
